# Is Kinesio taping effective for peripheral tissue perfusion in women with mild to moderate chronic venous insufficiency? A randomized controlled trial

**DOI:** 10.1101/2022.10.14.22281041

**Authors:** Maria Luiza Vieira Carvalho, Viviane De Menezes Caceres, Isabella de Oliveira Nascimento, Henrique Silveira Costa, Pedro Henrique Schedit Figueiredo, Vanessa Pereira Lima, Debora Pantuso Monteiro, Danielle Aparecida Gomes Pereira

## Abstract

**Objective:** To evaluate the acute effect of Kinesio Taping® on peripheral tissue perfusion in women with chronic venous insufficiency (CVI).

**Methods:** This randomized, double-blind controlled trial included 59 women with mild to moderate CVI. They were randomized to a control group (*n* = 23; 54.08 ± 9.04 years) and Kinesio Taping® group (KT) (*n* = 36; 55.87 ± 9.97 years). Near-infrared spectroscopy was positioned in the medial gastrocnemius muscle for assessment of resting tissue perfusion 48 h after the first day of evaluation and after placement of the Kinesio Taping® tape. The evaluation also consisted of performing movements of the plethysmography examination. To verify the comparisons of pre- and post-Kinesio Taping® data, the variation delta was used for analysis. Mann–Whitney U test was performed an an alpha of 5% was considered statistically significant.

**Results:** There wasn’
st a significant difference between groups regarding the peripheral tissue perfusion evaluation variables Peripheral Oxygen Saturation Difference – DELTA_SPO2: (KT Group 3.21 (0.84–3.62); Control 2.21 (1.59–4.83), p = 0.219) and Difference in deoxygenate hemoglobin values – DELTA_HHB (units) KT Group -0.62 (−2.14–0.67); Control Group -0.07 (−2.15–2.62) p = 0.238). Despite the lack of statistical significance, the KT group had a 785.7% greater drop in HHB values than the control group.

**Conclusions:** Acute use of Kinesio Taping® in women with CVI did not significantly alter tissue perfusion of calf muscles. However, it was possible to observe percentage differences in venous retention to be considered from a clinical point of view.

**Trial registration:** Brazilian Registry of Clinical Trials (REBEC) RBR-9bhbrp.

## Introduction

Chronic venous insufficiency (CVI) is characterized by persistent lower limb venous hypertension as a consequence of valvular incompetence, thrombosis, and a calf muscle pump function failure [1]. CVI has a prevalence of 17% in the U.S. adult population and is especially frequent in postmenopausal women because of a combination of various risk factors [2]. Among other symptoms, prolonged venous hypertension can lead to a perception of tiredness, heaviness, cramps, heat, or burning sensations on the skin or entire limb [3,4]. Individuals with CVI present with venous retention not only at rest but also during and after walking activities secondary to muscle pump dysfunction [5]. The severity of the disease has been related to reduced mobility of the talocrural joint, which worsens the gait of affected people and contributes to the emergence of venous ulcers [6]. Tissue damage in chronic venous insufficiency results from perivascular inflammation caused by a variety of cytokine mechanisms that weaken the usual dermal barrier against pathogens and allergens [7]. Lymphatic dysfunction, detected by means of nucleotide lymphangiography, is present in up to one third of cases of chronic venous insufficiency and may resolve with correction of the venous abnormalities [7].

The subjective complaints related to vein hypertension and reduced ankle range of ankle motion (ROAM) can be alleviated by compression therapy and an outpatient vascular exercise program [8]. However, treatment adherence, especially with compression therapy still low, and the main reasons include pain, discomfort, application difficulties, skin problems, uncomfortable footwear, and inadequate lifestyle advice from health care professional [8,9].

Kinesio Taping® (KT) is a therapeutic tool and has become increasingly popular in clinical practice for the treatment of orthopedic changes, pain, muscle changes, and edema [10]. In addition to these conditions, authors have proposed its use to reduce lymphatic and venous edema with promising results for symptoms of lower limb sensation, lameness, edema, cramps, pain, venous filling time, calf muscle power, and amount of extracellular fluid, the clinical severity of disease and physical function [11,12], and electromyographic activity of the gastrocnemius muscles [13]. However, little scientific evidence is available on the application of KT in people with vascular condition, and its usefulness in CVI has been poorly reported [11].

It is known that the homeostasis of interstitial fluid is critically important in the physiology and pathogenesis of tissues and is of therapeutic significance with important clinical implications [14]. The advocates of KT claim that convolution or mechanical deformation of the skin could lift epidermis away from the dermis altering the flow of interstitial fluid, blood, and lymph of the superficial tissues, which in turn might reduce swelling and pressure of pain receptors under the skin [15].

However, the precise action mechanism of KT is not fully explained, and to further understand its benefits in individuals with CVI, it is essential to evaluate acute tissue perfusion responses using this treatment technique. Support in the study of peripheral adaptations is offered by the use of near-infrared spectroscopy (NIRS) [16]. This technique noninvasively explores tissue microvascular hemodynamics, monitoring the local balance between oxygen delivery and consumption [16]. The NIRS technique allows exploration of a limited muscle region and offers a rapid study of the changes in perfusion, skeletal muscle blood flow, and mitochondrial respiratory function [14–16].

The present study aimed to evaluate the acute effect of KT on on peripheral tissue perfusion in women with CVI and compare with a placebo control group. The main hypothesis of the present study was that KT in the calf muscles could acutely improve cutaneous microvascular function and subsequent muscle oxidative metabolism in women with CVI.

## Methods

### Study Design

This study of acute effect of KT is part of a randomized, double-blind clinical trial. It was approved by the Research Ethics Committee of the Institution and registered at the Brazilian Registry of Clinical Trials (REBEC) - http://www.ensaiosclinicos.gov.br/rg/RBR-9bhbrp/.

The study was conducted between May 2017 and June 2018 and it was performed respecting the Declaration of Helsinki. In total, 63 volunteers were recruited; two were excluded owing to signs of deep vein thrombosis and heart failure and two owing to problems with data capture in NIRS (Fig 1).

**Fig 1.**
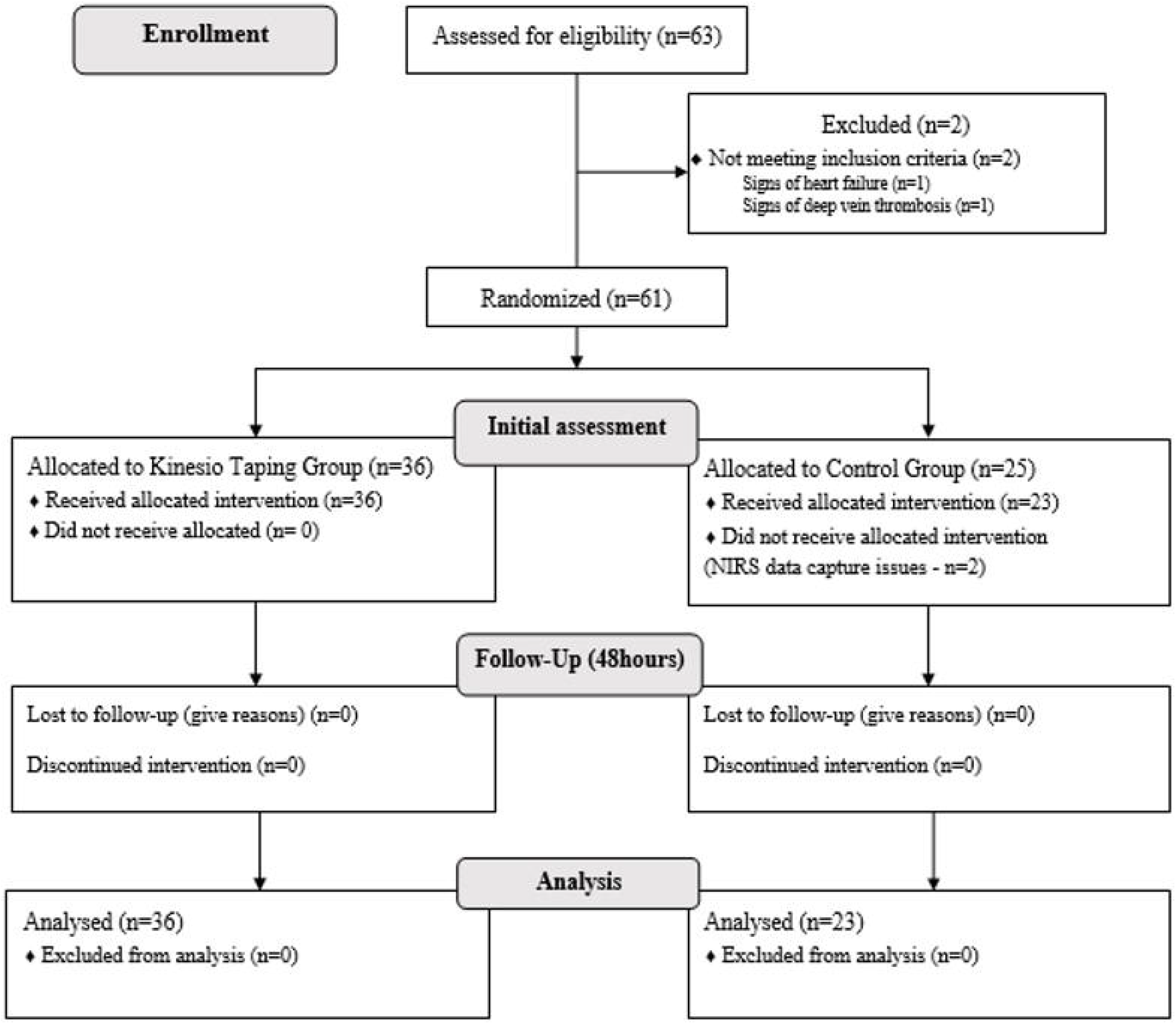
Flow chart of study. NIRS – Near Infrared Spectroscopy

Participants were randomized to the control group (GC) or the Kinesio Taping group (GKT) according to a list previously generated by the project coordinator using software available at www.randomization.com. The random allocation sequence was performed in blocks of four participants, allocation 1:1. Participants from both groups and evaluators were blinded in relation to GC and GKT allocation. Only the physical therapist who applied the KT tapes knew which group the participant belonged to.

### Participants

Sample selection was non-probabilistic, and all participants were informed about the risks and benefits of the study interventions, before signing the written informed consent form. The definitive sample size was calculated in GPower 3.1^®^ when the sample number reached 10 participants in each group. Considering the delta of the variables described for the study, the effect size found was 0.75, and the calculated *N* was 24 participants per group, for an alpha of 5% and a power of 80%.

From May 2017 to June 2018, women with CVI aged 30 to 79 years old, from Jenny de Andrade Faria Institute for Elderly and Women’s Health Care at the UFMG - Hospital das Clínicas, were enrolled in the study. The participants were included in the study according to the following inclusion criteria: being classified in stages C1, C2, or C3 of the Clinical Etiology Anatomy Pathophysiology Classification of Chronic Venous Disease (CEAP). The CEAP clinical item is classified from C0 to C6: C0 indicates the absence of clinical signs, C1 - telangiectasies or reticular veins, C2 - varicose veins, C3 - edema, C4 - skin changes and subcutaneous tissue, C5 - healed ulcer, and C6 - ulcer active [17]. The exclusion criteria on the study were: hormone replacement therapy, symptoms of intermittent claudication, lymphedema, thrombosis diagnosis for less than three months, contraindication to using KT as open wounds, experiences trauma, had edema from heart or kidney disease, had cancer, known KT tape allergy, and pregnancy. If the data captured by NIRS did not allow analysis, they were excluded.

### Experimental procedure

Two evaluations were performed for data collection, with an interval of 48 hours between assessments. The first day consisted of an initial assessment to collect demographic and clinical data such as age, sex, weight, height, body mass index, and analysis of disease severity according to CEAP of all participants included in the study (Fig 1). NIRS was positioned in the posterior calf region of the dominant leg (medially in the medial gastrocnemius muscle) to collect the Oxygen Saturation (StO_2_) and deoxyhemoglobin (HHB) data at rest for 3 min in the supine position. Subsequently, the volunteers performed the motion protocol of the plethysmography exam, which consisted of elevation of the lower limbs to a height of 15 cm for 5 min. After that, they were asked to move to the orthostatic position without support on the evaluated leg, until the stabilization of NIRS StO_2_ and HHB data, and were then requested to perform a bipodal plantar flexion. After plantar flexion, the participants were again instructed to keep the evaluated leg at rest and without support. After stabilization of the StO_2_ data, it was requested to perform ten plantar flexions in bipodal support. At the end of this procedure, the volunteer remained with the evaluated leg raised to stabilize the NIRS StO_2_ and HHB data [18].

On the second assessment day, the same protocol used in the first assessment to collect NIRS variables was performed on the second evaluation before and after KT placement. In GKT volunteers, the ribbons were placed in the prone position, the gastrocnemius muscles were stretched to measure the ribbons, and the “Y” cut was performed. Firstly, the leg was positioned in neutral, for fixation of the bases, without tension, in the proximal region of the popliteal fossa at the muscular origin, both in the medial and lateral regions. Subsequently, passive stretching of the triceps suralis muscle was performed by means of ankle dorsiflexion and KT placement with approximately 15% tension. In the end, the muscle stretching was removed, followed by the end of the application of the tensionless anchor. For participants of the CG, the KT was placed in the same places, but the necessary positions and tensions were not followed. NIRS variables were measured for 40 min before and after Kinesio Tex® [19] tape placement, after which, participants from both groups were asked to perform the movements of the air plethysmography protocol again.

### Outcome Measures

Muscle metabolism and related biomarkers were assessed by a continuous wave NIRS system (PortaMon System, ARTINIS MEDICAL SYSTEMS BV^®^). Near-infrared light (760 – 850 nm) propagating through biologic tissue is partly absorbed or scattered by the tissues and partly recollected by the detector; therefore, the intensity of the recollected light provides information about oxyhemoglobin (HbO_2_) and HHB concentrations. The equipment measures change in oxygenation in terms of HbO_2_ and HHB in real time. In addition, it calculates the StO_2_ index as a percentage, which reflects the average StO_2_ of muscle tissue. NIRS optodes with an interoptode distance of 3.5 cm, allowing light penetration of approximately 20 mm, were placed on the medial side of the calf (gastrocnemius muscles) and secured with tape.

The parameters considered were those from motion protocol of the plethysmography exam such as:

- Venous HHB volume (VV_HHB): the total venous filling volume of HHB after the individual has changed from supine to standing position.
- HHB venous filling index (IEV_HHB): the venous filling rate obtained by the ratio of 90% of VV_HHB to 90% of the filling time of VV_HHB.
- HHB value after plantar flexion (HHB_1FP)
- HHB ejection fraction (FE_HHB): calculated by the ratio of the HHB ejection volume (VE_HHB) to VV_HHB (FE_HHB = VE_HHB / VV_HHB).
- Venous retention fraction (FRET): the product of the ratio between the lowest HHB value after ten plantar flexions and the VV_HHB.
- Peripheral Oxygen Saturation Difference (DELTA_SPO_2_): result of subtracting SPO_2_ values before and after KT tape placement.
- Difference in deoxygenate hemoglobin values (DELTA_HHB): result of subtracting HHB values before and after KT tape placement.

### Statistical procedures

Statistical analysis of the data was performed by a researcher who was blinded to which group the sample volunteers belonged to. Data normality distribution was performed using the Shapiro-Wilk test and histograms. For descriptive analyses, categorical variables were presented as percentages (%), while numerical variables were described as means with their standard deviation (SD) or median with interquartile range (p25-p75), depending on their distribution. Independent t-test and chi-square test were used to compare baseline characteristics between groups. To verify comparisons of data before and after KT placement, the variation delta was used for analysis. Mann– Whitney U test was performed for peripheral tissue perfusion comparisons between the intervention and control groups. An alpha of 5% was considered statistically significant. The IBM Statistical Package for Social Sciences (SPSS) version 19.0 software was used for data analysis.

## Results

Out of the 63 women recruited for the study, 59 women with a mean age 54.78 ± 9.37 years met the inclusion criteria and were randomly assigned to the placebo control group (n: 23) or the experimental KT group (n: 36). The recruitment flowchart and follow-up of participants are summarized in Fig 1. Baseline demographic and clinical characteristics of the study sample were similar between groups for all variables (Table 1).

**Table 1.**
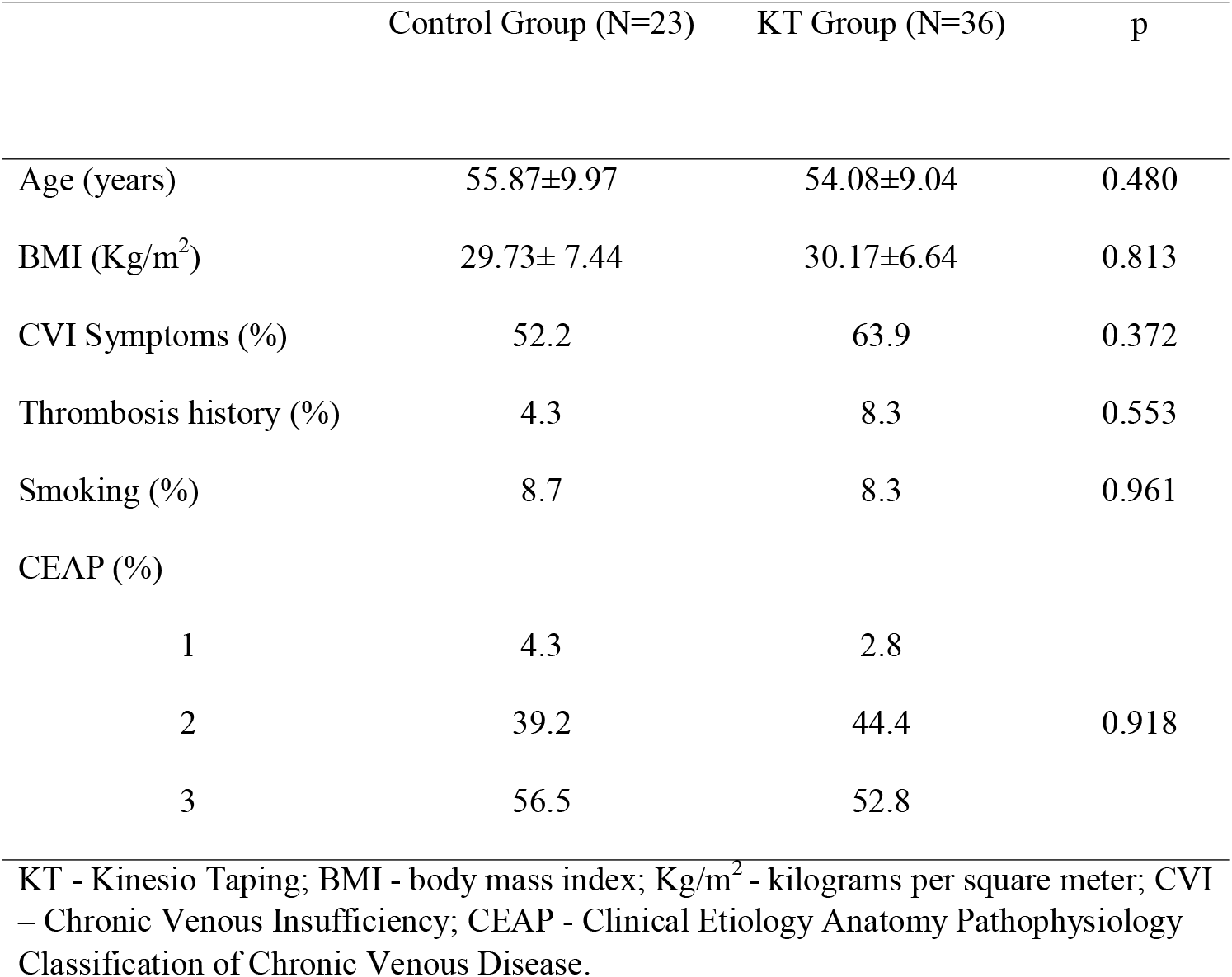
Baseline demographic and clinical characteristics of the sample.

|  | Control Group (N=23) | KT Group (N=36) | p |
| --- | --- | --- | --- |
| Age (years) | 55.87±9.97 | 54.08±9.04 | 0.480 |
| BMI (Kg/m<sup>2</sup>) | 29.73± 7.44 | 30.17±6.64 | 0.813 |
| CVI Symptoms (%) | 52.2 | 63.9 | 0.372 |
| Thrombosis history (%) | 4.3 | 8.3 | 0.553 |
| Smoking (%) | 8.7 | 8.3 | 0.961 |
| CEAP (%) |  |  |  |
| 1 | 4.3 | 2.8 |  |
| 2 | 39.2 | 44.4 | 0.918 |
| 3 | 56.5 | 52.8 |  |

KT - Kinesio Taping; BMI - body mass index; Kg/m<sup>2</sup> - kilograms per square meter; CVI – Chronic Venous Insufficiency; CEAP - Clinical Etiology Anatomy Pathophysiology Classification of Chronic Venous Disease.

Regarding the outcome variables of the peripheral tissue perfusion evaluation study, there were no significant posttreatment differences between groups (Table 2). Despite the absence of significant difference between groups, the KT group showed a 785.5% greater drop in HHB compared to the control group.

**Table 2.** Comparison of the variation of data obtained from NIRS in median and interquartile range (25-75) between KT group and control.

| Variables | Delta Control Group (N=23) | Delta KT Group (N=36) | Difference between groups (%) | p |
| --- | --- | --- | --- | --- |
| VV_HHB (units) | 1.03 (-8.91 – 6.36) | 0.80 (-4.44 – 0.80) | 22.8 | 0.384 |
| IEV_HHB (units /seconds) | 0.23 (0.06 – 0.69) | 0.33 (0.14 – 0.52) | 41.5 | 0.901 |
| HHB_1FP (units) | -0.19 (-10.26 – 5.32) | -0.30 (-5.00 – 5.39) | 57.9 | 0.465 |
| FE_HHB (%) | 19.59 (12.09 – 71.42) | 28.15 (9.98 – 63.93) | 43.7 | 0.852 |
| FRET (%) | 186.75 (97.16 – 318.53) | 142.79 (60.18 – 248.29) | 23.5 | 0.384 |
| DELTA_SPO<sub>2</sub> (units) | 2.21 (1.59 – 4.83) | 3.21 (0.84 – 3.62) | 31.0 | 0.219 |
| DELTA_HHB (units) | -0.07 (-2.15 – 2.62) | -0.62 (-2.14 – 0.67) | 785.7 | 0.238 |

Legend: KT - Kinesio Taping; VV\_HHB - Venous deoxyhemoglobin volume; IEV\_HHB - Deoxyhemoglobin venous filling index; HHB\_1FP -Deoxyhemoglobin value after plantar flexion; FE\_HHB - Deoxyhemoglobin ejection fraction; FRET - Venous retention fraction; DELTA\_SPO<sub>2</sub> - Peripheral Oxygen Saturation Difference; DELTA\_HHB - Difference in deoxyhemoglobin values.

Fig 2 shows the variation in HHB (DELTA_HHB) according to CEAP among women allocated to the control and KT groups. Although there was no statistically significant difference in DELTA_HHB between groups, there was a more evident graphic reduction in the median of this variable in women classified as C2 in the KT group. The median DELTA_HHb of women classified as CEAP 2 and allocated to the KT group (- 1.45 (−2.33-0.70)) dropped 56% more compared to its counterpart in the control group (- 0.63 (−2.90-2.14)).

**Fig 2.**
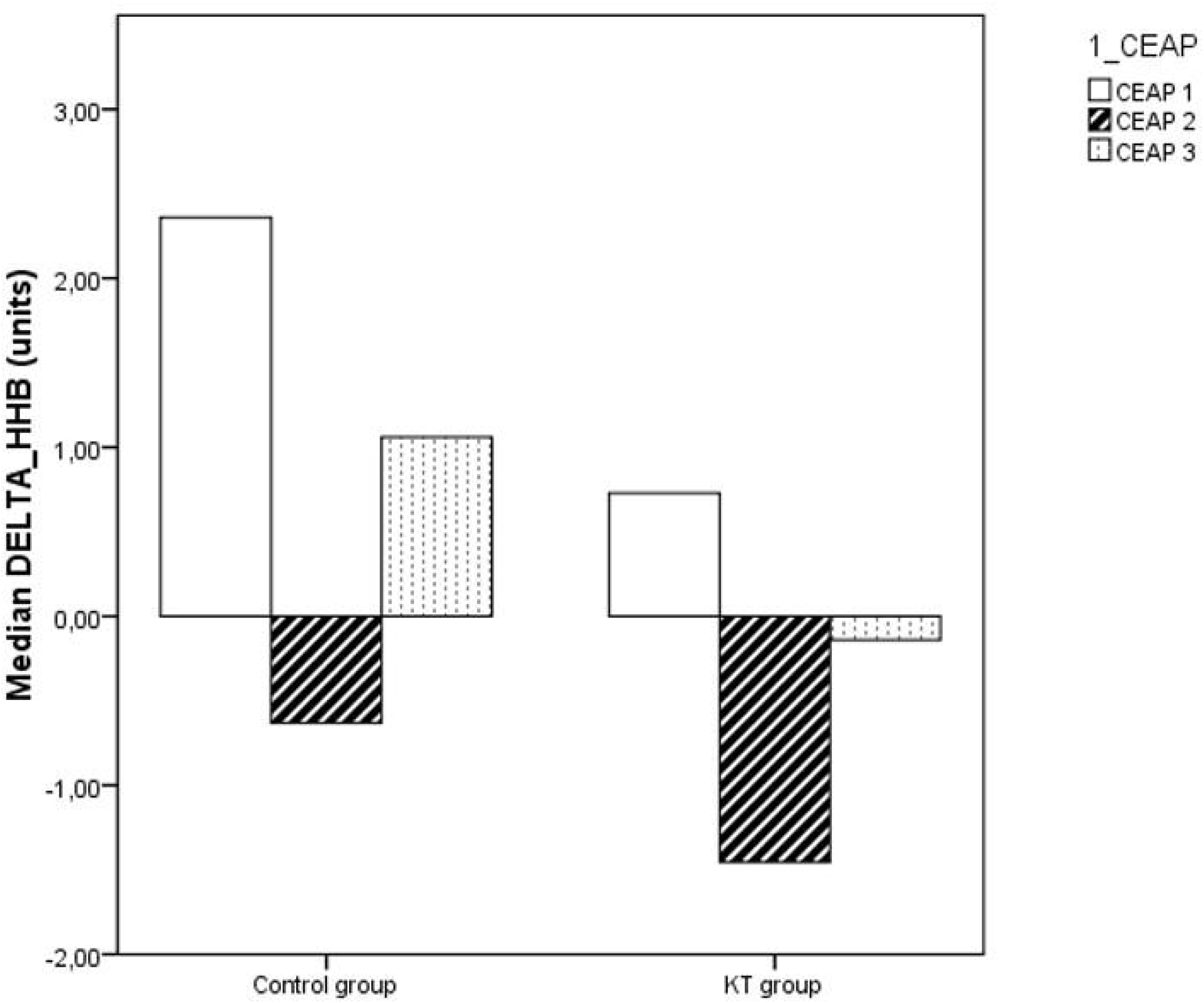
Variation in deoxygenated hemoglobin according to CEAP. DELTA_HHB: variation in deoxygenated hemoglobin; CEAP: Clinical Etiology Anatomy Pathophysiology Classification of Chronic Venous Disease.

## Discussion

In the present study, no statistically significant differences were found among variables that evaluated tissue perfusion between GKT and CG, in women with CVI, CEAP < 4 and overweight/obese. Despite this, it is important to highlight, from a clinical point of view, the differences in percentage values. GKT presented a 22.8% lower venous filling volume after KT tape placement than CG (placebo), indicating a change in venous system compliance. We could observe a 785.7% decrease in HHB value and a 31% increase in GKT SPO_2_ compared to CG. These alterations may suggest that clinical changes have occurred, since changes in the variables infer compliance and consequently modify venous hypertension and thus favor tissue oxygenation.

By analyzing the percentage changes in relation to the venous system filling index, GKT presented a 41.5% higher filling speed. In contrast, this group had HHB emptying at 57.9% greater plantar flexion, 43.7% better muscle pump power, and 23.5% lower venous retention. That is, although the venous system of GKT was more compliant, when there was muscle contraction, there was emptying of the vascular bed and less venous retention, indicating that using the tape may favor muscle activation and positively influence the venous system response.

We believe that it was not possible to verify a significant difference owing to the high variability of NIRS data, as well as the time of use of the tape. Aguilar-Ferrándiz et al (2013) [13] reported a protocol to use KT. A study was directed to responses regarding the electromyographic activity of the medial and lateral gastrocnemius muscles [13]. Throughout this study, the authors detected positive changes with increased muscle activity in women with CVI who used the tape three times a week for four weeks. Therefore, it can be inferred that chronic changes in muscle activation alter vascular responses, which in turn indirectly modify tissue perfusion, is directly dependent on the difference in intraluminal and tissue pressures. Therefore, the use of KT may be important in the treatment of venous stasis and may be used for secondary prevention of CVI.

The literature confirms that longer use of KT can also interfere with the symptoms and clinical variables of women with CVI, as shown in the study by Aguilar-Ferrándiz et al. (2013) [13], with reduced sensation of weight in the legs, lameness, venous vein, edema, and muscle cramps. In addition, changes in clinical parameters such as increased venous filling time and calf muscle power, assessed by photoplethysmography, occur. These data demonstrated that stimuli on skin surface caused by the placement of KT may cause changes in the venous system and perfusion [13]. However, the time of application, permanence of the tape, and form of evaluation appear to be relevant factors to be considered for its use.

In the present study, tissue perfusion was evaluated after 40 min of KT placement, and although there was no statistically significant change, the percentage concentration of HHB showed clinically relevant variation, which may contribute to the clinical practice for women with CVI. Tape usage time appears to be an important variable for more representative results, but this limitation is inherent in the proposed experimental design.

In the present study, the NIRS tissue perfusion assessment method was used; however, the variability of data associated with a small sample was a limiting factor for the results presented. Additionally, the sample consisted only of women with mild CVI severity. Studies should be conducted with a more heterogeneous sample, with greater disease severity.

## Conclusion

From the present study, it was possible to verify that acute use of KT in women with CVI could not alter tissue perfusion of the calf muscles significantly. However, it was possible to observe percentage differences in venous retention to be considered from a clinical point of view. Therefore, future studies are necessary to verify if long-term KT use could promote changes in tissue perfusion in women with CVI.

## Data Availability

All relevant data are within the manuscript and its Supporting Information files.

## Ethics Committee

approved by the Research Ethics Committee of the Federal University of Minas Gerais (CAAE 57708416.0.0000.5149).

## References

1. Eberhardt RT, Raffetto JD. Chronic venous insufficiency. Circulation. 2014;130: 333–346. doi:10.1161/CIRCULATIONAHA.113.006898

2. Bromen K, Pannier-Fischer F, Stang A, Rabe E, Bock E, Jöckel KH. [Should sex specific differences in venous diseases be explained by pregnancies and hormone intake?]. Gesundheitswesen. 2004;66: 170–174. doi:10.1055/S-2004-813019

3. Aldunate; JLCB, Isaac C, Ladeira PRS de, Carvalho VF, Ferreira MC. Venous ulcer in lower extremities. Rev Med. 2010;89: 158–163. doi:10.11606/ISSN.1679-9836.V89I3/4P158-163

4. Piazza G. Varicose veins. Circulation. 2014;130: 582–587. doi:10.1161/CIRCULATIONAHA.113.008331

5. Hosoi Y, Yasuhara H, Shigematsu H, Aramoto H, Komiyama T, Muto T. A new method for the assessment of venous insufficiency in primary varicose veins using near-infrared spectroscopy. J Vasc Surg. 1997;26: 53–60. doi:10.1016/S0741-5214(97)70147-2

6. Belczak CEQ, Godoy JMP de, Seidel AC, Silva JA, Junior GC, Belczak SQ. Assessing the influence of daily activities in the volumetry of inferior limbs via circumference measurement and water displacement volumetry. J Vasc Bras. 2004;3: 304–310.

7. Raju S, Neglén P. Chronic venous insufficiency and varicose veins. N Engl J Med. 2009;360: 2319–2327. doi:10.1056/NEJMCP0802444/SUPPL_FILE/NEJM_RAJU_2319SA1.PDF

8. Heinen MM, van der Vleuten C, de Rooij MJM, Uden CJT, Evers AWM, van Achterberg T. Physical activity and adherence to compression therapy in patients with venous leg ulcers. Arch Dermatol. 2007;143: 1283–1288. doi:10.1001/ARCHDERM.143.10.1283

9. Van Hecke A, Grypdonck M, Defloor T. A review of why patients with leg ulcers do not adhere to treatment. J Clin Nurs. 2009;18: 337–349. doi:10.1111/J.1365-2702.2008.02575.X

10. Gloviczki P, Comerota AJ, Dalsing MC, Eklof BG, Gillespie DL, Gloviczki ML, et al. The care of patients with varicose veins and associated chronic venous diseases: Clinical practice guidelines of the Society for Vascular Surgery and the American Venous Forum. J Vasc Surg. 2011;53: 2S–48S. doi:10.1016/J.JVS.2011.01.079

11. Aguilar-Ferrándiz ME, Castro-Sánchez AM, Matarán-Peñarrocha GA, Guisado-Barrilao R, García-Ríos MC, Moreno-Lorenzo C. A randomized controlled trial of a mixed Kinesio taping-compression technique on venous symptoms, pain, peripheral venous flow, clinical severity and overall health status in postmenopausal women with chronic venous insufficiency. Clin Rehabil. 2014;28: 69–81. doi:10.1177/0269215512469120/ASSET/IMAGES/LARGE/10.1177_0269215512469120-FIG1.JPEG

12. Aguilar-Ferrándiz ME, Moreno-Lorenzo C, Matarán-Peñarrocha GA, García- Muro F, García-Ríos MC, Castro-Sánchez AM. Effect of a mixed kinesio taping-compression technique on quality of life and clinical and gait parameters in postmenopausal women with chronic venous insufficiency: double-blinded, randomized controlled trial. Arch Phys Med Rehabil. 2014;95: 1229–1239. doi:10.1016/J.APMR.2014.03.024

13. Aguilar-Ferrándiz ME, Castro-Sánchez AM, Matarán-Peñarrocha GA, García-Muro F, Serge T, Moreno-Lorenzo C. Effects of kinesio taping on venous symptoms, bioelectrical activity of the gastrocnemius muscle, range of ankle motion, and quality of life in postmenopausal women with chronic venous insufficiency: a randomized controlled trial. Arch Phys Med Rehabil. 2013;94: 2315–2328. doi:10.1016/J.APMR.2013.05.016

14. Swartz MA, Fleury ME. Interstitial flow and its effects in soft tissues. Annu Rev Biomed Eng. 2007;9: 229–256. doi:10.1146/ANNUREV.BIOENG.9.060906.151850

15. Wiig H. Pathophysiology of tissue fluid accumulation in inflammation. J Physiol. 2011;589: 2945–2953. doi:10.1113/JPHYSIOL.2011.206136

16. Grassi B, Quaresima V. Near-infrared spectroscopy and skeletal muscle oxidative function in vivo in health and disease: a review from an exercise physiology perspective. J Biomed Opt. 2016;21: 91313. doi:10.1117/1.JBO.21.9.091313

17. Eklöf B, Rutherford RB, Bergan JJ, Carpentier PH, Gloviczki P, Kistner RL, et al. Revision of the CEAP classification for chronic venous disorders: Consensus statement. J Vasc Surg. 2004;40: 1248–1252. doi:10.1016/J.JVS.2004.09.027

18. Yamaki T, Nozaki M, Sakurai H, Takeuchi M, Soejima K, Kono T. The utility of quantitative calf muscle near-infrared spectroscopy in the follow-up of acute deep vein thrombosis. J Thromb Haemost. 2006;4: 800–806. doi:10.1111/J.1538-7836.2006.01859.X

19. Kase K, Tatsuyuki H, Tomoki O. Development of Kinesio Tape. Kinesio Taping Perfect Manual. Kinesio Taping Assoc. 1996;6: 117–188.

